# Diagnostic performance, implementation fidelity, and costs of the World Health Organization three-test HIV testing strategy in Malawi: a national retrospective evaluation

**DOI:** 10.64898/2026.08.30.26361753

**Authors:** Tiwonge Chimpandule, Hannock Tweya, Leah Goeke, Tobias Masina, Stephen Macheso, Nicola Low, Andreas Jahn, Jeffrey W. Imai-Eaton

**Affiliations:** Directorate of HIV, STIs and Viral Hepatitis, Ministry of Health, Lilongwe, Malawi; Institute of Social and Preventive Medicine, University of Bern, Bern, Switzerland; Graduate School for Cellular and Biomedical Sciences, University of Bern, Bern, Switzerland; Nyanja Health Research Institute, Lilongwe, Malawi; Quantitative Engineering Design (QED.ai), Lilongwe, Malawi; The Clinton Health Access Initiative (CHAI), Lilongwe, Malawi; Center for Communicable Disease Dynamics, Department of Epidemiology, Harvard T.H. Chan School of Public Health, Boston, MA, USA; MRC Centre for Global Infectious Disease Analysis, School of Public Health, Imperial College London, London, United Kingdom

**Keywords:** HIV testing, predictive value, three-test algorithm, Malawi, costs Word count: abstract 242, main text 3439

## Abstract

**Background:** In 2019, WHO recommended three consecutive reactive serological test results for HIV diagnosis to reduce false-positive diagnoses. Malawi changed from a two-test to a three-test strategy in 2022 as HIV test positivity declined. We assessed diagnostic performance, implementation fidelity, and costs.

**Methods:** We analysed national HIV testing data from Nov 1, 2022, to Oct 31, 2025. Using observed three-test classifications as the reference standard, we reconstructed classifications under the two-test strategy. We estimated positive predictive value (PPV), implementation fidelity, potential false-positive diagnoses prevented, incremental costs, and time to offset testing costs through avoided antiretroviral therapy expenditure.

**Results:** Among 9,885,599 encounters eligible for implementation-fidelity analysis, 99.98% followed a valid three-test pathway. The diagnostic-performance analysis included 9,862,908 encounters, of which 171,351 (1.7%) were classified HIV-positive and 9,138 (0.09%) were inconclusive. Under the two-test strategy, 1,209 inconclusive encounters with a T1+/T2+/T3− sequence would have been classified as HIV-positive. Retesting and reference-laboratory data indicated that 82.5% of these would subsequently be classified as HIV-negative, corresponding to 997 false-positive diagnoses prevented (10.3 per 100 000 three-test non-positive encounters; 95% CI 9.7–10.9). Retesting within 1–2 weeks was associated with the highest odds of potential false-positive classification (adjusted OR 39.37, 95% CrI 30.63–50.61). The incremental cost was US$471 per false-positive diagnosis averted and was offset within 7.30 years.

**Conclusions:** Malawi’s transition to a three-test HIV testing strategy prevented false-positive diagnoses and unnecessary antiretroviral therapy at modest cost, supporting broader adoption of WHO guidance in similar settings.

**Funding:** Gates Foundation.

## Introduction

Rapid diagnostic tests are used for HIV diagnosis in resource-limited settings because they are affordable, feasible to implement across all levels of health care, and provide clients and providers immediate feedback to guide linkage to further clinical treatment or prevention services.^1^ HIV rapid diagnostic tests are highly accurate, with specific products requiring ≥98% specificity to obtain WHO prequalification but are not perfect. Consequently, long-standing practice has been to require at least two consecutive reactive results with different tests to accurately diagnose HIV infection.^2^ As prevalence of a condition declines, even with highly accurate diagnostics, false-reactive results account for a greater proportion of all reactive results, reducing the positive predictive value of two-test strategies and increasing the risk of false-positive diagnoses.^3^ Such diagnoses can cause psychosocial harm, lead to unnecessary antiretroviral therapy (ART), waste limited resources, reduce confidence in HIV testing, and expose health systems and governments to legal consequences.^4^

In 2019, the World Health Organization (WHO) updated its guidance for HIV diagnosis in all settings to require reactive results from three consecutive rapid serological tests detecting antibodies to HIV. This recommendation replaced the previous strategy, under which two consecutive reactive tests were sufficient for diagnosis in settings with HIV prevalence of 5% or higher. The change aimed to maintain positive predictive value (PPV) above 99% as HIV test positivity declined.^1^ In modelling studies that assumed 98% specificity for each test, the minimum required for WHO prequalification, PPV fell below 99% with a two-test strategy when test positivity was below 5%, whereas a three-test strategy maintained high PPV at modest additional cost.^2^ Despite this guidance, adoption has remained incomplete: by December 2021, only 45% (21 of 47) of countries in the WHO African Region implemented a three-test strategy.^5^ Reported barriers included algorithm verification, procurement, budget planning, additional test-kit requirements, staff workload, and possible delays in ART initiation.^5–7^ However, direct empirical evidence on the diagnostic performance, implementation fidelity, and costs of the three-test strategy under routine national implementation remains limited.

In Malawi, HIV test positivity declined from 4.6% in 2016 to 2.1% in 2022.^8^ The country transitioned from a two-test to a three-test algorithm in November 2022 with phased rollout.^9^ To inform decisions on three-test adoption in other low- and middle-income countries with declining HIV positivity, we estimated the PPV of the two-test strategy, potential false-positive diagnoses prevented, implementation fidelity, factors associated with potential false-positive classification, and incremental costs.

## Methods

### Study design, setting, and data sources

We conducted a diagnostic evaluation of the national HIV programme in Malawi, from Nov 1, 2022, to Oct 31, 2025. We used results from the observed three-test HIV testing strategy and compared them with the outcome that would have been delivered if the two-test strategy had still been in place. We also assessed implementation fidelity, factors associated with potential false-positive diagnosis, and incremental diagnostic commodity costs. The unit of analysis was the testing encounter; individuals who tested more than once during the study period contributed multiple encounters. Testing used rapid diagnostic tests on finger-prick capillary whole blood^8^ and was performed primarily by trained lay health workers.^10^ The three-test algorithm was introduced alongside ScanForm™ (QED.ai, Lilongwe, Malawi), a digital data-capture system in which testing providers use smartphones to photograph completed HIV-testing register pages. Optical character recognition digitises the handwritten entries into encounter-level records.^9^

By Oct 31, 2025, the dataset covered 863 of Malawi’s 867 HIV-testing facilities (99.5%) and community-based testing recorded through these facilities, including outreach services and established community testing points documented under a supervising facility. We report this study in accordance with STARD 2015,^11^ where applicable (appendix S5).

### Inclusion and exclusion criteria

All HIV-testing encounters with at least one rapid diagnostic test result recorded in facility or community testing registers and available in ScanForm™ were eligible. All eligible encounters contributed to the assessment of implementation fidelity, defined as whether the recorded assays followed the prescribed testing sequence and whether the recorded final HIV result was consistent with the assay results. Analyses of diagnostic performance excluded encounters with a testing sequence that deviated from the national algorithm or a compliant sequence with an inconsistent final result. We also excluded encounters that did not reach a definitive classification because the required testing sequence was not completed; these encounters included testing in community settings with a reactive result from the first rapid test, which required referral to a facility and encounters in health facilities where the session ended before all required assays were completed.

### HIV diagnostic test algorithm and analytic approach

The rapid diagnostic tests used in the Malawi HIV diagnostic algorithm were, in sequence, Determine™ HIV-1/2 (test 1, [T1]; Abbott Diagnostics, Chiba, Japan), Uni-Gold HIV (T2; Trinity Biotech, Bray, Ireland), and SD Bioline HIV-1/2 3.0 (T3; Standard Diagnostics, Yongin-si, Republic of Korea). WHO prequalification assessments reported sensitivities of 100%, 99.76%, and 100%, and specificities of 98.9%, 99.9%, and 99.9%, respectively.^12–14^

Under the three-test algorithm, three consecutive reactive (+) results (T1+/T2+/T3+) were required for an HIV-positive diagnosis (figure S1). A non-reactive (-) T1 result was classified as HIV-negative and testing ended. A discordant T1+/T2− result triggered repeat testing with T1. A T1+/T2+/T3− result was classified as inconclusive and required follow-up retesting after two weeks.^8^

We reconstructed the classification of HIV infection from each encounter under the former two-test strategy using T1 and T2 results only (figure S2). We used the observed three-test result as the reference standard; T1− encounters were classified as HIV-negative and T1+/T2+ encounters as HIV-positive. T1+/T2− results would have prompted same-visit repetition by another provider under the two-test algorithm; 0% of these encounters remained discordant after repeat testing in the two-test period and were classified as inconclusive in the simulation.^15^

The primary outcome was potential false-positive classification under the simulated two-test strategy, defined as a T1+/T2+/T3− encounter that was reactive after T1 and T2 but not confirmed by T3. The three-test confirmation proportion among encounters classified as positive under the simulated two-test strategy was calculated as T1+/T2+/T3+ encounters divided by all T1+/T2+ encounters; its complement represented the potential false-positive proportion. Because HIV status was not independently verified, the three-test strategy was treated as the best available but imperfect reference standard.^16^ Apparent specificity relative to this strategy was the proportion of encounters classified as HIV-negative or inconclusive under the three-test strategy that were also classified as non-positive under the simulated two-test strategy. The corresponding 2×2 classification table and algebraic derivations are in appendix S2 (tables S3 and S4). Potential false-positive frequency was reported as an absolute number and per 100 000 three-test non-positive encounters, with exact binomial 95% CIs. Sensitivity was not estimated because no independent reference standard was available to identify clients falsely classified as HIV-negative.

### Factors associated with a potential false-positive diagnosis under a simulated two-test strategy

We used client and encounter characteristics recorded in the HIV testing register to identify factors associated with potential false-positive classification under the simulated two-test strategy, including sex, age, access mode, time since the previous HIV test, and antiretroviral-use history. Access modes were provider-initiated testing and counselling, voluntary counselling and testing, mobile or outreach testing, social-network testing, and index testing. Index testing was defined as active when a health-care provider initiated tracing and testing of contacts of a person diagnosed with HIV, and passive when contacts presented after referral by the index case. Antiretroviral use comprised pre-exposure prophylaxis, post-exposure prophylaxis, or ART. Analyses were restricted to encounters classified as HIV-negative or inconclusive under the three-test algorithm; only these could have been classified as HIV-positive by the two-test strategy but not confirmed by T3. The outcome was a T1+/T2+/T3− sequence. We fitted a hierarchical binomial logistic regression model with a facility-level random intercept, diffuse Gaussian priors on coefficients (mean 0, precision 0.001), and a log-gamma prior on the random-intercept precision. Estimates are reported as posterior median ORs with 95% CrIs. Missing sex and age were retained as ‘Not recorded’; missing time was included in ‘Never tested or missing’. No imputation was performed.

### Estimating the proportion of false-positive results among inconclusive results after applying the three-test algorithm

Clients with a T1+/T2+/T3− result were informed that the result was inconclusive and asked to return after two weeks for repeat rapid diagnostic testing. If the repeat result remained inconclusive, a dried-blood-spot sample was sent to the national reference laboratory for confirmatory testing

We estimated the proportion of T1+/T2+/T3− results later found to be HIV-negative using facility retesting among clients retested within three months and inconclusive samples referred to the national reference laboratory. Reference laboratory data were available from October 2023 to March 2025. Because these data sources could not be linked at the individual level, the estimated proportions were applied at the aggregate level to the observed number of T1+/T2+/T3− outcomes (appendix S1; figure S3). We assessed the plausibility of this aggregate approach by comparing estimates stratified by sex and age group, and by examining facility-level agreement between expected persistent inconclusive cases and samples processed by the reference laboratory (appendix S3; figures S4–S6).

### Diagnostic commodity cost analysis

We compared the cost from a health-system payer perspective of implementing the three-test strategy versus the two-test strategy in 2025 US dollars (US$).^17^ Incremental diagnostic and personnel costs were compared with avoided lifetime ART care costs (commodities, personnel, overhead, supervision, shared consumables).

Diagnostic commodity unit costs were obtained from the Global Fund pooled procurement reference prices for 2025.^18^ Personnel costs were estimated from a previously-published Malawi HIV-testing episode cost analysis^19^ and applied only to the additional testing steps required by the three-test algorithm. Annual ART care costs were estimated at US$73 per patient-year by combining 2025 Global Fund commodity prices^18^ with delivery, supervision, and overhead costs from a provider-costing study in rural Malawi.^19^ Avoided ART care costs were accrued over an expected 30-year remaining life expectancy for someone initiating ART, and discounted at 3% annually, consistent with previous analyses.^3^

We conducted sensitivity analyses varying two parameters: the proportion of test kits not used for testing individual clients, including kits lost, expired, or used for provider training and quality control; and the proportion of T1+/T2− encounters that remained discordant after same-session repeat testing. Detailed costing inputs and assumptions are in appendix S4 (tables S5–S7).

Analyses were conducted in R version 4.5.1 (R Foundation for Statistical Computing, Vienna, Austria). Bayesian logistic regression was implemented via R-INLA package (version 25.10.19).

### Ethical approval

Ethical approval was obtained from the Malawi National Health Sciences Research Committee (protocol 23/12/4275).

### Role of the funding source

The funders had no role in study design, data collection, analysis, interpretation, or the decision to submit for publication.

## Results

Between Nov 1, 2022, and Oct 31, 2025, 11,597,932 HIV testing encounters were recorded in Malawi. Per the phased rollout, 1,712,333 (14.8%) encounters occurred before the respective facilities transitioned to the three-test algorithm and individual-level record reporting, and individual-level records were available for 9,885,599 (85.2%) encounters from 863 of Malawi’s 867 HIV-testing facilities (99.5%), including community testing conducted through these facilities. These 9,885,599 encounters were included in the implementation-fidelity analysis (figure 1).

**Figure 1:**
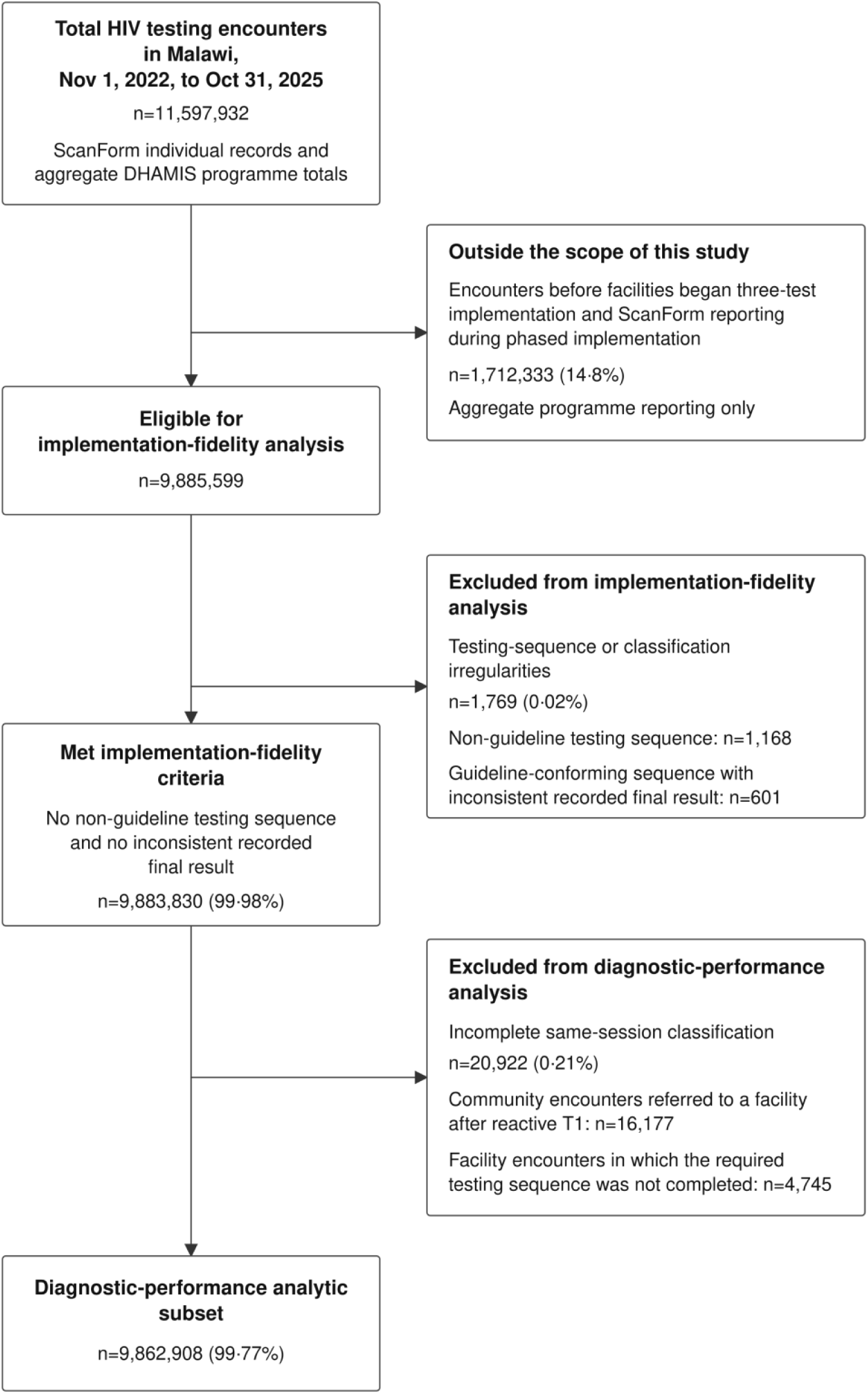
Inclusion and exclusion of HIV testing encounters for the implementation-fidelity and diagnostic-performance analyses. Percentages in the two lowest boxes are of the 9 885 599 encounters eligible for the implementation-fidelity analysis. Data are HIV testing encounters recorded in Malawi between Nov 1, 2022, and Oct 31, 2025.

Implementation fidelity, defined as adhering to a guideline-conforming testing sequence with a consistent final recorded result, was 99.98% (9,883,830/9,885,599 encounters). The 1,769 encounters that did not meet these criteria comprised 1,168 with non-guideline testing sequences and 601 with guideline-conforming sequences but an inconsistent recorded final result. These irregularities could not be verified but 477 could have affected the recorded classification; 458 could have resulted in a false-positive classification and 19 in a false-negative classification (appendix tables S1 and S2).

Among the 9,883,830 encounters that met the implementation-fidelity criteria, 20,922 (0.21%) did not have a final HIV status classification and were excluded from the diagnostic-performance analysis. These comprised 16,177 community encounters with a reactive T1+ result and referred to facility testing, in accordance with guidelines, and 4,745 facility encounters in which the required testing sequence was not completed. The diagnostic-performance analysis therefore comprised 9,862,908 encounters with a complete three-test classification of HIV-negative, HIV-positive, or inconclusive (figure 1).

Most encounters were among female clients (6,713,860 [68.1%]) and clients aged 15–49 years (8,306,004 [84.2%]). Provider-initiated testing and counselling accounted for 7,582,706 (76.9%) encounters and voluntary counselling and testing for 1,263,016 (12.8%). More than half of encounters involved clients whose last HIV test was more than 6 months previously (5,369,268 [54.4%]), whereas 2,648,093 (26.9%) involved clients who had never previously tested or whose testing history was missing. Most encounters involved clients reporting no previous antiretroviral use (9,557,125 [96.9%]; table 2).

Among the 9,862,908 encounters in the diagnostic-performance analysis, 9,682,419 (98.2%) were classified as HIV-negative, 171,351 (1.7%) as HIV-positive, and 9,138 (0.09%) as inconclusive under the three-test algorithm (figure 2). The HIV-negative classifications comprised 9,681,154 (99.99%) encounters with a non-reactive T1 result and 1,265 (0.013%) with a non-reactive repeat T1 result after initial T1+/T2– discordance. The inconclusive classifications comprised 7,929 encounters with a reactive repeat T1+ result after initial T1+/T2– discordance and 1,209 with a T1+/T2+/T3− sequence.

**Figure 2:**
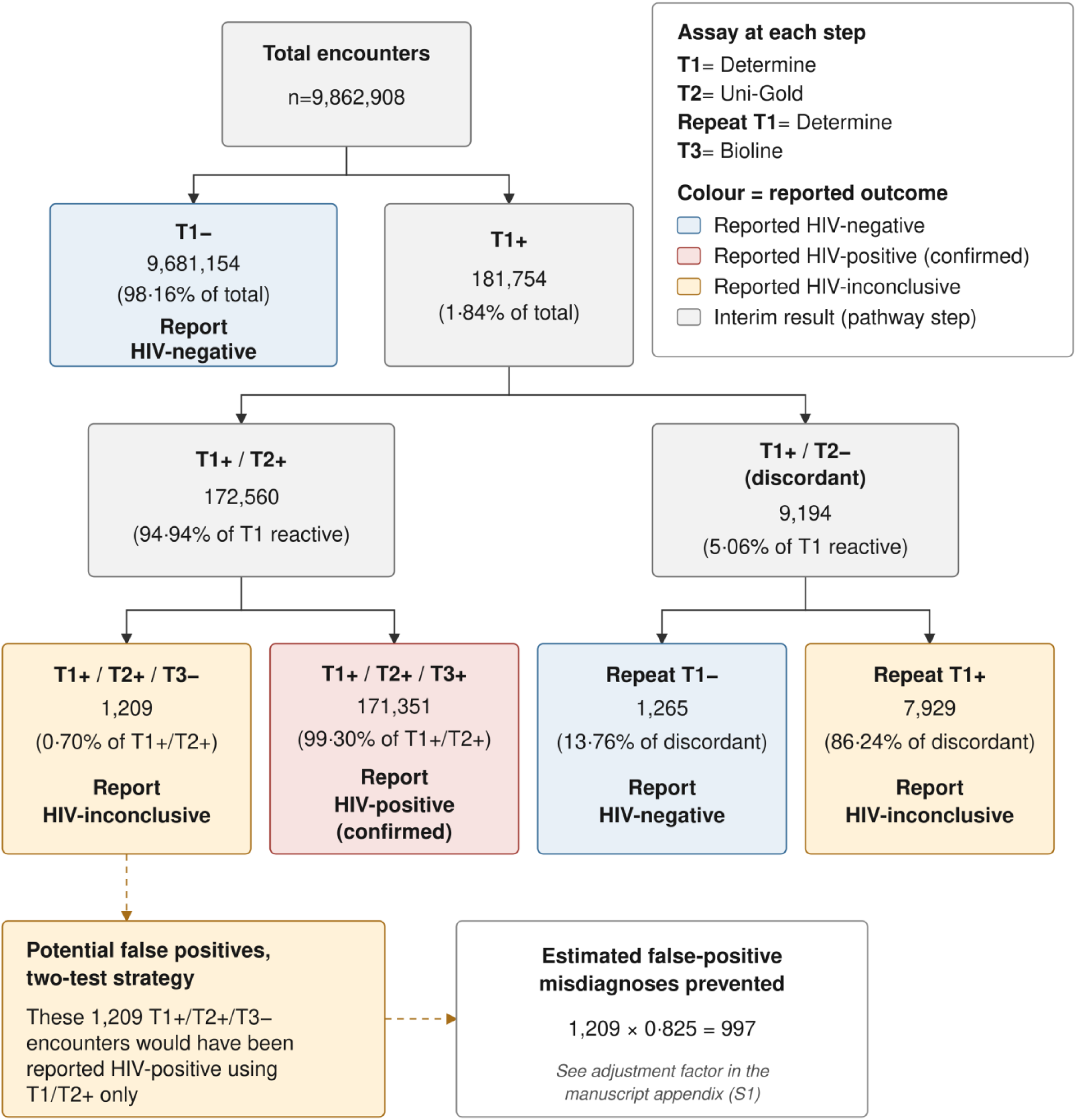
Flow of HIV testing encounters through the three-test algorithm and misdiagnoses prevented by the confirmatory third test. Encounters are the diagnostic-performance analytic subset defined in figure 1. Percentages are of the immediate parent category. Box colour denotes the reported outcome (blue, HIV-negative; red, HIV-positive [confirmed]; amber, HIV-inconclusive; grey, interim step). Three consecutive reactive results (T1+/T2+/T3+) are required for an HIV-positive diagnosis. Assays: T1 and repeat T1, Determine HIV-1/2; T2, Uni-Gold HIV; T3, Bioline HIV-1/2. Derivation of the 0.825 adjustment factor is shown in appendix S1.

Under the two-test strategy, the 1,209 encounters with a T1+/T2+/T3− sequence would have been classified as HIV-positive. Under the implemented three-test strategy, these encounters were classified as inconclusive and referred for follow-up retesting. Facility retesting and national reference-laboratory data indicated that 82.5% of these results would subsequently be classified as HIV-negative (appendices S1 and S3). The proportion was similar by sex (82.0% and 83.0%) and across age groups (81.0% to 84.0%). Applying this proportion yielded an estimated 997 potential false-positive diagnoses prevented, equivalent to 10.3 (95% CI 9.7–10.9) per 100 000 encounters classified as HIV-negative or inconclusive under the three-test algorithm (table 1). The simulated two-test strategy had a PPV of 99.30% (95% CI 99.26–99.34).

**Table 1:**
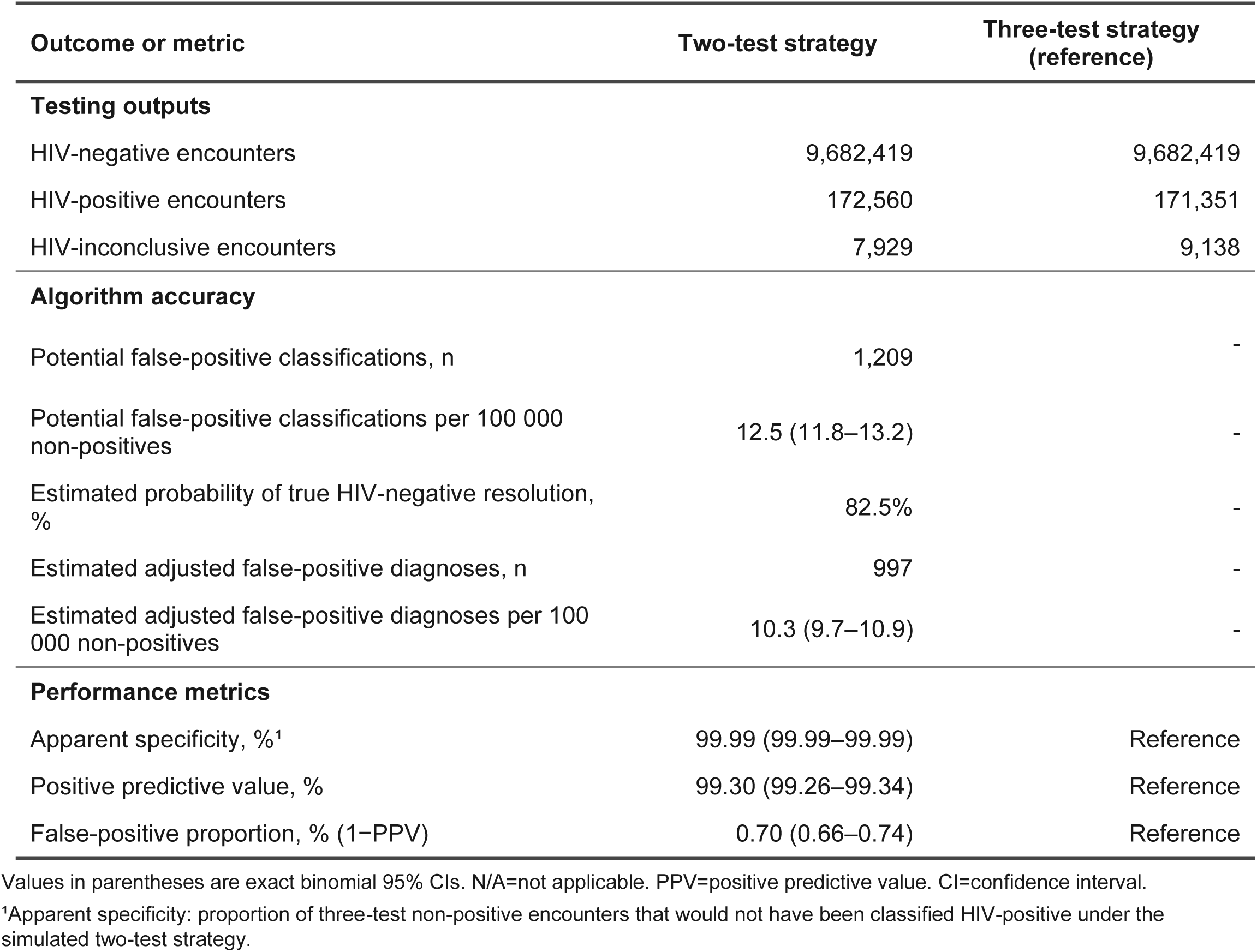
Diagnostic performance and potential false-positive classifications under the simulated two-test strategy.

The odds of a potential false-positive classification under the simulated two-test strategy varied substantially across subgroups (table 2). In adjusted analyses, clients retesting within 1–2 weeks had approximately 39 times higher odds than those last tested more than 6 months previously (adjusted OR 39.37, 95% CrI 30.63–50.61). Odds were also higher among clients tested through passive index testing (5.27, 4.08–6.80) or voluntary counselling and testing (1.85, 1.60–2.15). Compared with adults aged 25–49 years, odds were higher among adults aged 50 years or older (1.85, 1.51–2.27) and lower among those aged 15–24 years (0.76, 0.66–0.86), 5–9 years (0.43, 0.25–0.76), and 10–14 years (0.53, 0.32–0.89). Odds were also lower among females than males (0.87, 0.77–0.99). Reported ART use was associated with higher odds than no antiretroviral use (4.41, 1.38–14.05), although the CrI was wide and only three potential false-positive classifications occurred in this group.

**Table 2:** HIV tests, HIV-positive diagnoses, and potential false-positive HIV classifications under the simulated two-test strategy, by subgroup.

| Covariate | Tested (N) | HIV-positive (N) | Positivity, % (95% CI) | Potential FP (N) | FP proportion, % (1–PPV) | Non-positive denominator (N) | FP per 100 000 non-positives (95% CI) | Crude OR (95% CrI) | Adjusted OR (95% CrI) |
| --- | --- | --- | --- | --- | --- | --- | --- | --- | --- |
| Overall | 9,862,908 | 171,351 | 1.74 (1.73–1.75) | 1,209 | 0.70 (0.66–0.74) | 9,691,557 | 12.5 (11.8–13.2) | N/A | N/A |
| Sex |  |  |  |  |  |  |  |  |  |
| Male | 3,144,592 | 66,860 | 2.13 (2.11–2.14) | 441 | 0.66 (0.60–0.72) | 3,077,732 | 14.3 (13.0–15.7) | 1.00 (ref) | 1.00 (ref) |
| Female | 6,713,860 | 104,491 | 1.56 (1.55–1.57) | 768 | 0.73 (0.68–0.78) | 6,609,369 | 11.6 (10.8–12.5) | 0.79 (0.70–0.89) | 0.87 (0.77–0.99) |
| Not recorded /missing | 4,456 | 0 | 0.00 (0.00–0.08) | 0 | N/A | 4,456 | 0.0 (0.0–82.8) | N/A | N/A |
| Age group (years) |  |  |  |  |  |  |  |  |  |
| 0–4 | 565,576 | 3,599 | 0.64 (0.62–0.66) | 63 | 1.72 (1.32–2.20) | 561,977 | 11.2 (8.6–14.3) | 1.03 (0.79–1.35) | 0.91 (0.68–1.21) |
| 5–9 | 285,301 | 1,443 | 0.51 (0.48–0.53) | 14 | 0.96 (0.53–1.61) | 283,858 | 4.9 (2.7–8.3) | 0.47 (0.27–0.80) | 0.43 (0.25–0.76) |
| 10–14 | 256,006 | 1,758 | 0.69 (0.66–0.72) | 16 | 0.90 (0.52–1.46) | 254,248 | 6.3 (3.6–10.2) | 0.56 (0.34–0.92) | 0.53 (0.32–0.89) |
| 15–24 | 4,032,045 | 34,212 | 0.85 (0.84–0.86) | 413 | 1.19 (1.08–1.31) | 3,997,833 | 10.3 (9.4–11.4) | 0.73 (0.64–0.83) | 0.76 (0.66–0.86) |
| 25–49 | 4,273,959 | 114,726 | 2.68 (2.67–2.70) | 586 | 0.51 (0.47–0.55) | 4,159,233 | 14.1 (13.0–15.3) | 1.00 (ref) | 1.00 (ref) |
| 50+ | 445,106 | 15,611 | 3.51 (3.45–3.56) | 117 | 0.74 (0.62–0.89) | 429,495 | 27.2 (22.5–32.6) | 1.96 (1.60–2.39) | 1.85 (1.51–2.27) |
| Not recorded /Missing | 4,915 | 2 | 0.04 (0.00–0.15) | 0 | 0.00 (0.00–84.19) | 4,913 | 0.0 (0.0–75.1) | N/A | N/A |
| Access mode |  |  |  |  |  |  |  |  |  |
| Provider-initiated testing and counselling | 7,582,706 | 94,969 | 1.25 (1.24–1.26) | 817 | 0.85 (0.80–0.91) | 7,487,737 | 10.9 (10.2–11.7) | 1.00 (ref) | 1.00 (ref) |
| Voluntary counselling and testing | 1,263,016 | 33,733 | 2.67 (2.64–2.70) | 303 | 0.89 (0.79–1.00) | 1,229,283 | 24.6 (22.0–27.6) | 1.85 (1.60–2.13) | 1.85 (1.60–2.15) |
| Mobile | 386,375 | 1,302 | 0.34 (0.32–0.36) | 4 | 0.31 (0.08–0.78) | 385,073 | 1.0 (0.3–2.7) | 0.08 (0.03–0.23) | 0.11 (0.04–0.29) |
| Outreach | 115,344 | 267 | 0.23 (0.20–0.26) | 4 | 1.48 (0.40–3.74) | 115,077 | 3.5 (0.9–8.9) | 0.26 (0.10–0.69) | 0.33 (0.12–0.88) |
| Social network strategy | 79,082 | 2,979 | 3.77 (3.64–3.90) | 2 | 0.07 (0.01–0.24) | 76,103 | 2.6 (0.3–9.5) | 0.20 (0.05–0.81) | 0.26 (0.06–1.03) |
| Active index | 258,864 | 4,061 | 1.57 (1.52–1.62) | 4 | 0.10 (0.03–0.25) | 254,803 | 1.6 (0.4–4.0) | 0.15 (0.06–0.40) | 0.22 (0.08–0.58) |
| Passive index | 177,521 | 34,040 | 19.18 (18.99–19.36) | 75 | 0.22 (0.17–0.28) | 143,481 | 52.3 (41.1–65.5) | 5.10 (3.99–6.51) | 5.27 (4.08–6.80) |
| Time since last HIV test |  |  |  |  |  |  |  |  |  |
| Same day | 39,218 | 7,922 | 20.20 (19.80–20.60) | 17 | 0.21 (0.12–0.34) | 31,296 | 54.3 (31.6–87.0) | 8.56 (4.90–14.97) | 10.01 (5.76–17.41) |
| 1–6 days | 24,670 | 3,739 | 15.16 (14.71–15.61) | 13 | 0.35 (0.18–0.59) | 20,931 | 62.1 (33.1–106.2) | 5.30 (3.05–9.20) | 6.26 (3.60–10.88) |
| 1–2 weeks | 18,069 | 720 | 3.98 (3.70–4.28) | 72 | 9.09 (7.18–11.31) | 17,349 | 415.0 (324.9–522.4) | 34.56 (26.93–44.34) | 39.37 (30.63–50.61) |
| 2–3 weeks | 14,873 | 231 | 1.55 (1.36–1.76) | 7 | 2.94 (1.19–5.97) | 14,642 | 47.8 (19.2–98.5) | 4.00 (1.89–8.44) | 5.12 (2.42–10.83) |
| 3–4 weeks | 328,296 | 2,178 | 0.66 (0.64–0.69) | 68 | 3.03 (2.36–3.82) | 326,118 | 20.9 (16.2–26.4) | 1.80 (1.40–2.32) | 2.04 (1.57–2.64) |
| 1–2 months | 350,063 | 1,776 | 0.51 (0.48–0.53) | 49 | 2.68 (1.99–3.53) | 348,287 | 14.1 (10.4–18.6) | 1.23 (0.92–1.65) | 1.49 (1.10–2.00) |
| 3–6 months | 1,070,358 | 7,871 | 0.74 (0.72–0.75) | 102 | 1.28 (1.04–1.55) | 1,062,487 | 9.6 (7.8–11.7) | 0.81 (0.65–1.00) | 0.96 (0.77–1.18) |
| >6 months | 5,369,268 | 120,320 | 2.24 (2.23–2.25) | 628 | 0.52 (0.48–0.56) | 5,248,948 | 12.0 (11.0–12.9) | 1.00 (ref) | 1.00 (ref) |
| Never tested or missing | 2,648,093 | 26,594 | 1.00 (0.99–1.02) | 253 | 0.94 (0.83–1.07) | 2,621,499 | 9.7 (8.5–10.9) | 0.89 (0.77–1.04) | 1.13 (0.96–1.32) |
| Antiretroviral use history |  |  |  |  |  |  |  |  |  |
| None | 9,557,125 | 163,395 | 1.71 (1.70–1.72) | 1,163 | 0.71 (0.67–0.75) | 9,393,730 | 12.4 (11.7–13.1) | 1.00 (ref) | 1.00 (ref) |
| Post-exposure prophylaxis | 48,454 | 858 | 1.77 (1.66–1.89) | 10 | 1.15 (0.55–2.11) | 47,596 | 21.0 (10.1–38.6) | 1.55 (0.83–2.91) | 1.09 (0.58–2.05) |
| Pre-exposure prophylaxis | 247,723 | 1,532 | 0.62 (0.59–0.65) | 33 | 2.11 (1.46–2.95) | 246,191 | 13.4 (9.2–18.8) | 1.32 (0.92–1.88) | 0.94 (0.65–1.36) |
| Antiretroviral therapy | 9,606 | 5,566 | 57.94 (56.95–58.93) | 3 | 0.05 (0.01–0.16) | 4,040 | 74.3 (15.3–216.9) | 4.63 (1.48–14.53) | 4.41 (1.38–14.05) |

Tested, HIV-positive, and positivity use all encounters. The non-positive denominator for potential-FP rates and models comprised three-test negative or inconclusive encounters (n=9,691,557). Potential false-positive classifications were T1+/T2+/T3– encounters under the simulated two-test strategy (n=1,209). Rates are per 100 000 non-positive encounters with exact binomial 95% CIs. ORs are posterior medians (95% CrIs) from INLA models with a facility random intercept; adjusted models include all covariates shown. Missing sex and age are shown as 'Not recorded'; missing time and legacy 2–3-month records are included in 'Never tested or missing' for comparability with appendix table S4.
N/A=not applicable. FP=false-positive. PPV=positive predictive value. OR=odds ratio. CI=confidence interval. CrI=credible interval.

Estimated diagnostic testing costs were US$8,661,952 under the simulated two-test strategy and US$9,131,867 under the observed three-test strategy, an incremental cost of US$469,915 (5.4% higher; table 3). Personnel was the largest contributor to the incremental cost, accounting for US$222,315 (47.3%), followed by additional diagnostic commodities at US$164,899 (35.1%). Other delivery and overhead costs contributed US$51,133 (10.9%), and supervision and monitoring and evaluation costs contributed US$31,569 (6.7%). Thus, non-commodity costs collectively accounted for 64.9% of the incremental cost. The incremental cost per false-positive diagnosis averted was US$471.

**Table 3:** Cost of the three-test algorithm compared with the two-test algorithm.

| Metric | Two-test strategy | Three-test strategy | Incremental (three-test – two-test) |
| --- | --- | --- | --- |
| **Scale of impact** |  |  |  |
| Clients analysed | 9,862,908 | 9,862,908 | Same population |
| Potential false-positive classifications | 1,209 | N/A | –1,209 |
| False-positive diagnoses averted | N/A | 997 | +997 |
| **Test kit utilisation** |  |  |  |
| Total test kits used | 10,549,125 | 10,773,655 | +224,530 |
| Client-use kits | 10,070,405 | 10,226,416 | +156,011 |
| Non-client or wastage kits | 478,720 | 547,239 | +68,519 |
| **Diagnostic testing costs (2025 US\$)** | | | |
| Total diagnostic testing cost | \$8,661,952 | \$9,131,867 | +\$469,915 |
| Diagnostic commodity cost | \$8,661,952 | \$8,826,851 | +\$164,899 |
| Additional personnel cost | \$0 | \$222,315 | +\$222,315 |
| Additional supervision/M&E cost | \$0 | \$31,569 | +\$31,569 |
| Additional other delivery/overhead cost | \$0 | \$51,133 | +\$51,133 |
| Cost per false-positive diagnosis averted | - | - | \$471 |
| **ART care costs avoided (2025 US\$)** | | | |
| Annual ART care cost per person | - | \$73 | \$73 |
| Annual ART care costs avoided | - | \$72,682 | \$72,682 |
| Discounted lifetime ART care cost per person | - | \$1,428 | \$1,428 |
| Discounted lifetime ART care costs avoided | - | \$1,424,606 | \$1,424,606 |
| **Cost offset** |  |  |  |
| Undiscounted time to cost offset, years | - | 6.47 | 6.47 |
| Discounted time to cost offset, years | - | 7.30 | 7.30 |
| Net cost offset (ART costs avoided minus diagnostic cost) | - | \$954,691 saved | \$954,691 saved |
| **Standardised outcomes (per 100 000 tested)** |  |  |  |
| False-positive diagnoses averted | - | 10.11 | 10.11 |
| Incremental testing cost | - | +\$4,764 | +\$4,764 |
| Discounted ART care costs avoided | - | \$14,444 | \$14,444 |
| Net cost offset | - | \$9,680 saved | \$9,680 saved |

All costs are in 2025 US dollars. N/A=not applicable to that strategy column. Hybrid Malawi-specific programme or provider cost scenario; base-case wastage 4% (T1) and 43% (T2 or T3); persistent discordance after same-session repeat 40%. False-positive diagnoses averted=potential false-positive classifications (T1+/T2+/T3–) × 0.825 adjustment factor.
Diagnostic cost components follow the grouped cost categories defined in appendix S3 (table S5): Commodity (test kits and commodities), Delivery (personnel and health-worker time), Management (supervision, audits, and M&E), and Other delivery or overhead (other supplies, utilities, capital, and overheads).
Full costing methods, sources, and sensitivity analyses are in appendix S3 (tables S5 and S6).

Discounted lifetime ART care costs avoided across 997 false-positive diagnoses averted were US$1,424,606, giving a net cost offset of US$954,691 over the lifetime horizon. The undiscounted and discounted times to cost offset were 6.47 years and 7.30 years, respectively. Across sensitivity analyses, incremental diagnostic costs ranged from US$437,679 to US$489,887, and the discounted time to cost offset ranged from 6.74 years to 7.65 years (figure S7). In all scenarios, discounted ART care costs avoided exceeded incremental diagnostic testing costs.

## Discussion

Implementing the WHO-recommended three-test strategy was estimated to have prevented 997 potential false-positive diagnoses during the first three years of programme implementation, corresponding to around 10 misdiagnoses prevented per 100 000 three-test non-positive encounters. Implementation of the three-test strategy had high fidelity, with 99.98% of encounters compliant. The incremental cost was US$471 per potential false-positive diagnosis averted and was offset within 7.30 years by avoided ART care expenditure.

A major strength of this study is the national dataset, enabled by digitisation of routine HIV-testing records from 99.5% of Malawi’s HIV-testing facilities. These data reflect implementation in routine programme conditions, largely delivered by trained lay providers. Most previous PPV estimates rely on survey-based evaluations^20–22^ or on modelling with assumed assay specificity.^2^ We combined clinical and laboratory data sources to estimate the proportion of inconclusive results subsequently expected to be determined to be HIV-negative. Applying the same recorded test results to the observed three-test and simulated two-test strategies enabled direct comparison of the two algorithms. Individual-level data also enabled retrospective identification of 1,769 encounters with data-quality irregularities, including 477 that could have led to an incorrect result. This identification is essential for monitoring implementation and directing mentorship to sites and providers where quality improvement is needed. While these potential classification errors were only identified retrospectively and anonymously in our study, in future the real-time digitised records could be leveraged to proactively monitor, contact, and recall clients where errors may have occurred to reverify their HIV status.

The 99.30% PPV estimated for the simulated two-test strategy is broadly comparable with estimates from routine national programmes and population-based surveys in similar settings. Across 14 population-based HIV impact assessment surveys, Patel and colleagues reported a pooled adult PPV of 99.4% in the ten countries using a two-test algorithm without a tie-breaker and 98.8% in the four using a tie-breaker; the 2015–16 Malawi survey reported a PPV of 96.5%.^21^ In a similar household survey in Nigeria, PPV was 94.5% at 1.4% prevalence, which the authors attributed to low prevalence and tester-related error under survey conditions.^20^ Eaton and colleagues reported substantial improvement in Malawi’s HIV testing programme between 2014 and 2016: the proportion of initial HIV-positive diagnoses subsequently found to be HIV-negative on verification retesting decreased from 7% to 1%, which they attributed to implementation of WHO-recommended testing strategies, updated national guidelines, and tester retraining. In 2017, verification testing records showed that approximately 1% of initial HIV-positive results were subsequently classified as HIV-negative, corresponding to a programme PPV of approximately 99%.^3^ Our estimated PPV of 99.30% is therefore broadly consistent with the more recent programme performance reported in Malawi. It is higher than the 97.7% predicted at 1.7% positivity by a modelling study assuming that each assay attained the WHO-prequalification minimum specificity of 98%,^2^ but lower than the approximately 99.91% expected if the assays performed at their published specificities and errors were independent.^12,13^ The difference from this theoretical value might reflect lower specificity under routine field conditions, correlated false reactivity between assays, or undetected procedural and documentation errors.^22,23,24^

Several limitations should be acknowledged. The observed three-test result was a programme reference standard rather than an independent measure of true HIV status, and outcomes were unavailable for clients with inconclusive results who did not return. The estimate of 997 potential false-positive diagnoses prevented relied on an aggregate adjustment factor derived from facility-retesting and reference-laboratory data. Associations from the regression analysis are subject to confounding by unmeasured client or provider factors not captured in routine data, and documentation errors could have caused residual misclassification. Finally, the cost analysis was restricted to the health-system payer perspective and did not include one-time provider-training costs, operational effects on staff workload and client waiting times, or wider patient and societal costs.

Our study suggests that the benefits of the transition to a three-test algorithm outweigh the harms. Concerns have been raised that three-test algorithms cause harmful delay in ART initiation.^25^ Our findings, based on analysis of reference laboratory retesting, suggest that about four in five inconclusive results from the three-test algorithm would eventually be determined to be HIV-negative; the third test therefore prevented unnecessary lifelong ART. In the remainder, who would be found to be living with HIV, ART would be delayed by the interval to follow-up retesting (typically 4–6 weeks). The cost-saving conclusion is robust: net programme savings of US$955,000 and offset within 7.30 years held across all sensitivity analyses. Additional test use was modest, with T3 volume equivalent to 1.75% of T1 volume. The incremental cost of US$471 per false-positive diagnosis averted was comparable with the estimated US$460 for verification testing in Malawi^2^; both add a confirmation step before determining lifelong treatment.

Our study has implications for practice and research. As of December 2021, most countries in the WHO African Region continued to recommend a two-test strategy in national policy.⁵ Malawi’s experience shows that high implementation fidelity is achievable at national scale within a programme delivered mainly by lay health workers.^10^ At 1.7% positivity, a two-test PPV of 99.30% still generated 997 preventable misdiagnoses over three years. In programmes with lower algorithm PPV, this burden would be substantially higher. The risk of a potential false-positive result under the simulated two-test strategy varied substantially across testing contexts and client groups, and may reflect nonspecific serological reactivity, immune activation, weak-line interpretation, specimen handling, or correlated assay errors.^23,24^ These mechanisms warrant further investigation. In the subgroups at higher risk, additional verification or targeted provider training could reduce misdiagnosis in countries that still use a two-test algorithm.

This national empirical evaluation shows that Malawi’s transition to the WHO three-test HIV testing strategy prevented false-positive diagnoses and unnecessary lifelong ART at modest additional cost, supporting broader adoption of WHO guidance in similar settings.

## Data Availability

De-identified encounter-level data used in this study will be made available to researchers with a legitimate scientific purpose, following approval of a data-access agreement by the Malawi Ministry of Health. Analysis scripts will be made available alongside the data. Requests should be submitted to the Ministry of Health through the Department of HIV and AIDS with a brief description of the proposed analysis; access decisions will be made by the Malawi Ministry of Health under a signed data-access agreement.

## Acknowledgements

TC and JWI-E were supported by the Bill & Melinda Gates Foundation (INV-005576). We thank the Malawi Ministry of Health for access to the national HIV testing data, and the health workers who recorded it. The content is solely the responsibility of the authors and does not necessarily represent the views of the funder.

Under the grant conditions of the Foundation, a Creative Commons Attribution 4.0 License has already been assigned to the Author Accepted Manuscript version that might arise from this submission.

## Declaration of interests

Hannock Tweya and Leah Goeke work with QED.ai, which implemented the ScanForm system used by the national programme.

## Use of AI-assisted technology in manuscript preparation

Tiwonge Chimpandule used artificial intelligence tools (Claude, Anthropic) to assist with language editing and reference formatting during preparation of this manuscript. He reviewed and edited all AI-assisted outputs as needed. All authors reviewed and approved the final manuscript and take full responsibility for its content. No generative AI tool was used for data collection, data analysis, or generation of the research findings.

## Authorship contributions

TC: Conceptualisation, Methodology, Formal analysis, Visualisation, Writing - original draft, Writing - review & editing

HT: Methodology, review & editing

LG: Data curation, Resources, Writing - review & editing TM: Resources, review & editing

SM: Resources, review & editing

NL: Supervision, Methodology, review & editing

AJ: Supervision, Methodology, Data curation, Writing - review & editing

JWI-E: Supervision, Methodology, Data curation, Resources, Writing - review & editing

## Notes

### Author Declarations

National Health Sciences Research Committee (NHSRC),Malawi

