## Supplementary Material for "Diagnostic performance, implementation fidelity, and costs of the World Health Organization three-test HIV testing strategy in Malawi: a national retrospective evaluation"

### Appendix S1. Supplementary Results Tables and Figures

**Table S1**- Excluded data-quality irregularities: compliant testing sequences with discordant recorded outcomes

| Pathway | Setting | Allowed outcome(s) | Recorded outcome | n | % within inconsistent results | Severity | Interpretation |
|---|---|---|---|---|---|---|---|
| ***High risk of inaccurate adverse outcome delivered to client*** | | | | | | | |
| T1+ | Facility | Initial positive | Positive | 375 | 62·4% | High | Conclusive outcome recorded despite non-conclusive pathway |
| T1+ | Community | Initial positive | Positive | 29 | 4·8% | High | |
| T1+ | Facility | Initial positive | Positive | 28 | 4·7% | High | |
| T1+, T2+, T3+ | Facility | Positive | Negative | 19 | 3·2% | High | Reactive sequence classified as negative |
| T1− | Facility | Negative | Positive | 4 | 0·7% | High | Potential false-positive classification |
| T1+ | Facility | Initial positive | Exposed infant | 2 | 0·3% | High | |
| T1+ | Facility | Initial positive | Negative | 1 | 0·2% | High | Potential false-negative classification |
| | | | **Sub Total High severity** | **458** | **76·2%** | | |
| ***High risk of inaccurate adverse outcome delivered to client*** | | | | | | | |
| T1+, T2−, T1R− | Facility | Negative | Inconclusive | 71 | 11·8% | Low | Non-conclusive outcome (retest required) |
| T1+, T2+, T3+ | Facility | Positive | Inconclusive | 36 | 6·0% | Low | |
| T1+ | Facility | Initial positive | Inconclusive | 36 | 6·0% | Low | |
| | | | **Sub Total low severity** | **143** | **23·8%** | | |
| | | | **Total** | **601** | **100·0%** | | |

**Table S2**- Excluded data-quality irregularities: non-compliant testing sequences, by pattern, recorded outcome, and risk classification

| Pattern | Recorded outcome | n | % non-compliant | Risk | Violation | Interpretation |
|---|---|---|---|---|---|---|
| ***High risk of inaccurate adverse outcome delivered to client*** | | | | | | |
| T1+, T2−, T3−, T1R− | Positive | 11 | 0·9% | High | Discordant sequence with positive outcome | Potential false-positive classification |
| T1+, T2+, T3+, T1R− | Negative | 4 | 0·3% | High | Three reactive tests followed by negative outcome | Incorrect negative |
| T1+, T2−, T3−, T1R− | Positive | 2 | 0·2% | High | Discordant sequence with positive outcome | Potential false-positive |
| T1+, T2+, T3−, T1R+ | Positive | 1 | 0·1% | High | Repeat T1R after a valid sequence | Potential false positive |
| T1+, T2−, T3−, T1R− | Positive | 1 | 0·1% | High | Repeated discordant testing | Inconsistent classification |
| **Subtotal, high risk** | ·· | **19** | **1·6%** | | | |
| ***Low risk of inaccurate adverse outcome delivered to client*** | | | | | | |
| T1−, T2+, T3+ | Positive | 511 | 43·8% | Low | T1 missing or non-reactive before confirmatory tests | Procedural or documentation error |
| T1+, T2+, T3+, T1R+ | Positive | 271 | 23·2% | Low | Repeat T1 after three reactive tests | Unnecessary repeat testing |
| T1+, T2+, T3−, T1R+ | Inconclusive | 63 | 5·4% | Low | Discordant T3 followed by repeat testing | Non-conclusive outcome |
| T1+, T2−, T3−, T1R+ | Inconclusive | 42 | 3·6% | Low | Repeat testing after discordant sequence | Non-conclusive outcome |
| T1+, T2−, T3+, T1R+ | Inconclusive | 34 | 2·9% | Low | Non-sequential testing | Non-conclusive outcome |
| T1−, T2+, T3+ | Positive retest | 29 | 2·5% | Low | T1 missing or non-reactive before confirmatory tests | Procedural or documentation error |
| T1+, T2+ | Positive initial | 24 | 2·1% | Low | Incomplete algorithm | Non-conclusive classification |
| T1−, T2−, T1R+ | Inconclusive | 22 | 1·9% | Low | Repeat testing after inconsistent sequence | Non-conclusive outcome |
| T1+, T2+, T3+, T1R+ | Positive retest | 16 | 1·4% | Low | Repeat T1 after three reactive tests | Unnecessary repeat testing |
| T1+, T2+, T3+, T1R− | Inconclusive | 14 | 1·2% | Low | Reactive sequence recorded as non-conclusive | Non-conclusive outcome |
| Complex discordant | Negative | 13 | 1·1% | Low | Non-compliant sequence | Inconsistent classification |
| T1 missing, T2+, T3+ | Positive retest | 11 | 0·9% | Low | Missing T1 with reactive confirmatory tests | Procedural or documentation error |
| T1+, T2+ | Not concluded | 10 | 0·9% | Low | Incomplete algorithm | Non-conclusive classification |

| Pattern | Recorded outcome | n | % non-compliant | Risk | Violation | Interpretation |
|---|---|---|---|---|---|---|
| T1 missing, T2+, T3+ | Positive | 10 | 0·9% | Low | Missing T1 with reactive confirmatory tests | Procedural or documentation error |
| Other minor patterns | Mixed | 79 | 6·8% | Low | Various deviations | Other non-compliant outcomes |
| **Subtotal, low risk** | | **1149** | **98·4%** | | | |
| **Total** | | **1168** | **100·0%** | | | |

Violation describes how each sequence deviates from the WHO-recommended serial testing algorithm. Pos retest=positive retest; pos initial=initial positive pending confirmation. T1=Determine HIV-1/2; T2=Uni-Gold HIV; T3=SD Bioline HIV-1/2 3.0; T1R=repeat test 1.

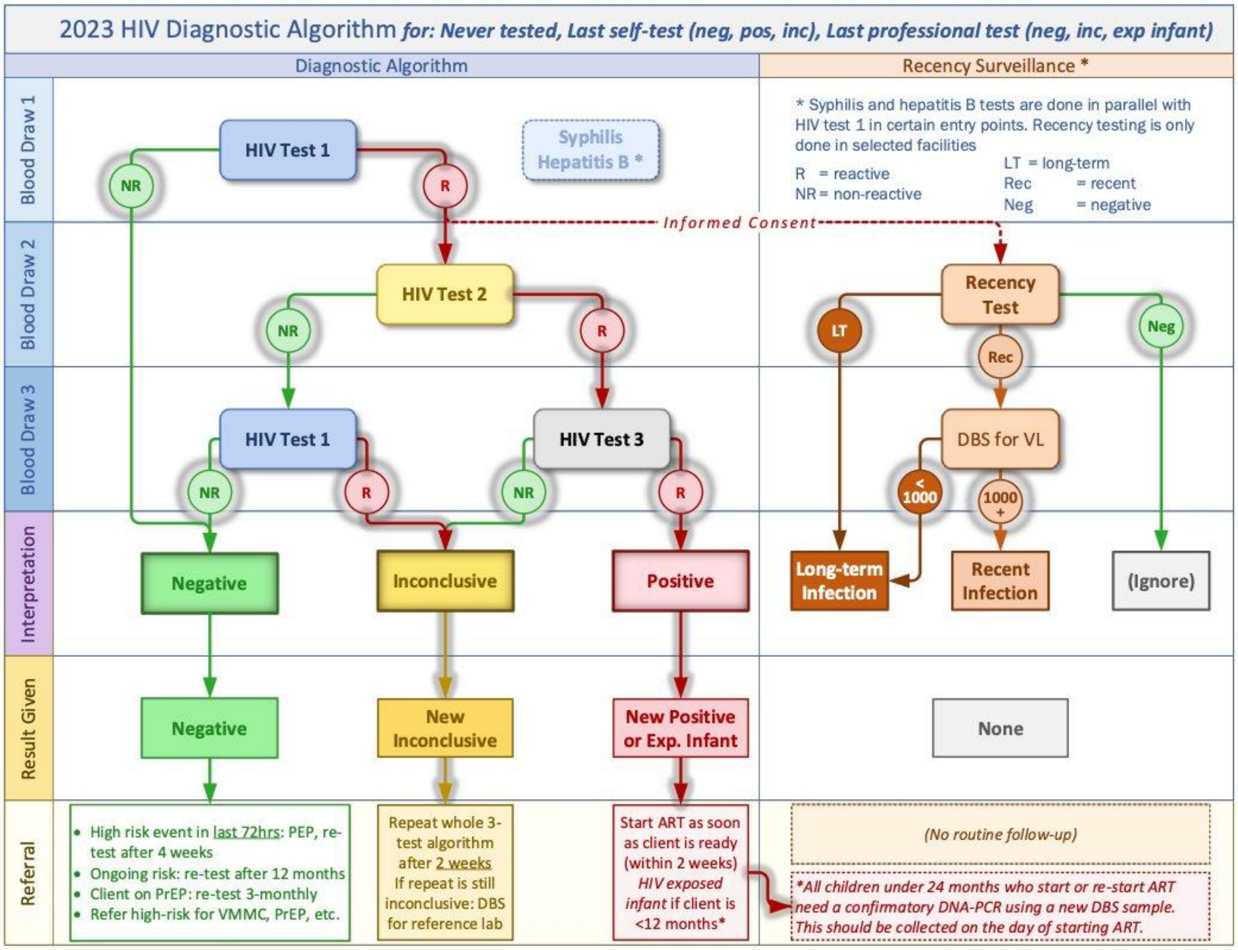

**Figure S 1.** Three-test HIV testing algorithm (Malawi, implemented since November 2022)

Reproduced from: Ministry of Health, Republic of Malawi. HIV, Syphilis and Hepatitis B Integrated Rapid Testing and Counselling Guidelines and Standard Operating Procedures. 1st edition. Lilongwe: Ministry of Health; 2023 (algorithm shown on page 61). T1=Determine HIV-1/2; T2=Uni-Gold HIV; T3=SD Bioline HIV-1/2 3·0. R=reactive; NR=non-reactive. DBS=dry blood spot; VL=viral load; LT=long-term; Rec=recent; Neg=negative

**Figure S1** presents the national three-test HIV diagnostic algorithm implemented in Malawi from November 2022,[1] in accordance with WHO 2019 HIV testing guidelines.[2] The algorithm requires three consecutive reactive results (T1+/T2+/T3+) for a confirmed HIV-positive diagnosis.

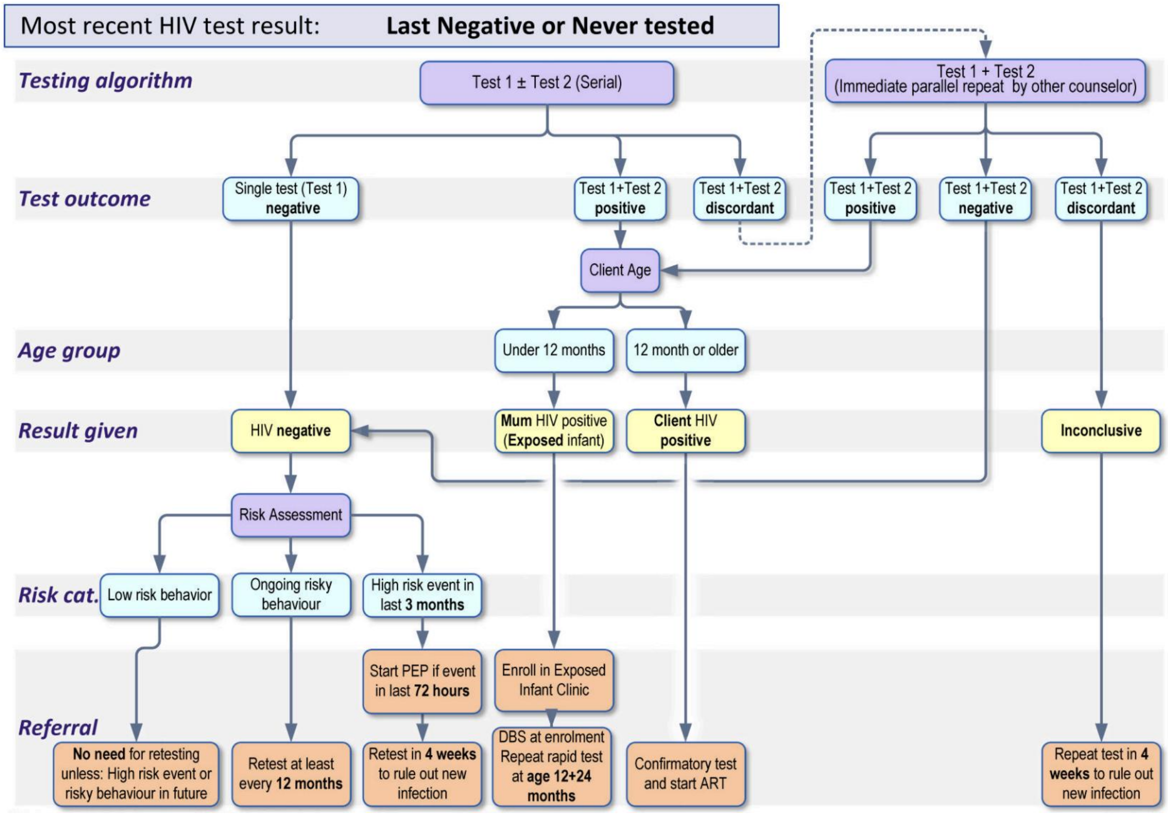

**Figure S2** Two-test HIV diagnostic algorithm (Malawi, pre-November 2022 counterfactual).

Reproduced from: Ministry of Health, Republic of Malawi. HIV Testing Services Guidelines. Lilongwe: Ministry of Health; 2016 (algorithm shown on page 56). T1=Determine HIV-1/2; T2=Uni-Gold

**Figure S2** above presents the former two-test HIV diagnostic algorithm used in Malawi before November 2022,[3] simulated as the counterfactual strategy in this analysis. Under this algorithm, two consecutive reactive results (T1+/T2+) were sufficient for a confirmed HIV-positive diagnosis.

### Appendix S2. Estimating the proportion of T1+/T2+/T3− results classified as HIV-negative

#### S2.1 Rationale

The primary analysis identified 1209 HIV testing encounters with a T1+/T2+/T3− result pattern. Under the simulated two-test algorithm, these encounters would have been classified HIV-positive; under the three-test algorithm they were classified inconclusive and referred for follow-up testing. Some of these inconclusive tests referred for follow-up testing may ultimately be determined as truly HIV positive.

Routine HIV testing services (HTS) data were encounter-based and did not allow deterministic linkage of individual clients across repeat testing visits and reference laboratory confirmation. Consequently, the final HIV classification of each T1+/T2+/T3− encounter could not be directly observed.

#### S2.2 Data sources and derivation

To estimate the proportion of these encounters that were ultimately classified as HIV-negative, an empirical probability was derived using two programme cohorts representing sequential stages in the national HIV testing pathway:

- HTS retesting cohort: individuals who previously received an inconclusive HIV result and returned for repeat testing within three months.
- Reference laboratory cohort: individuals with persistent inconclusive results referred for confirmatory testing at the national reference laboratory (October 2023 to March 2025).

The probability that a T1+/T2+/T3− encounter was ultimately classified as HIV-negative was estimated as:

#### S2.3 Adjustment factor estimates

Among 2487 individuals returning for repeat testing after an inconclusive result, outcomes were 42·4% HIV-negative on repeat testing, 14·7% HIV-positive on repeat testing, and 42·9% persistently inconclusive (Figure S3). Among 1121 samples referred for laboratory confirmation, outcomes were 93·6% HIV-negative and 6·4% HIV-positive on laboratory testing.

Substituting these empirical estimates: $0{\cdot}424 + (0{\cdot}429 \times 0{\cdot}936) = 0{\cdot}825$. Thus, an estimated 82·5% of T1+/T2+/T3− encounters would ultimately be determined to be HIV-negative. Applying this probability to the 1209 observed T1+/T2+/T3− encounters yielded an estimated 997 potential false-positive diagnoses prevented by the third test.

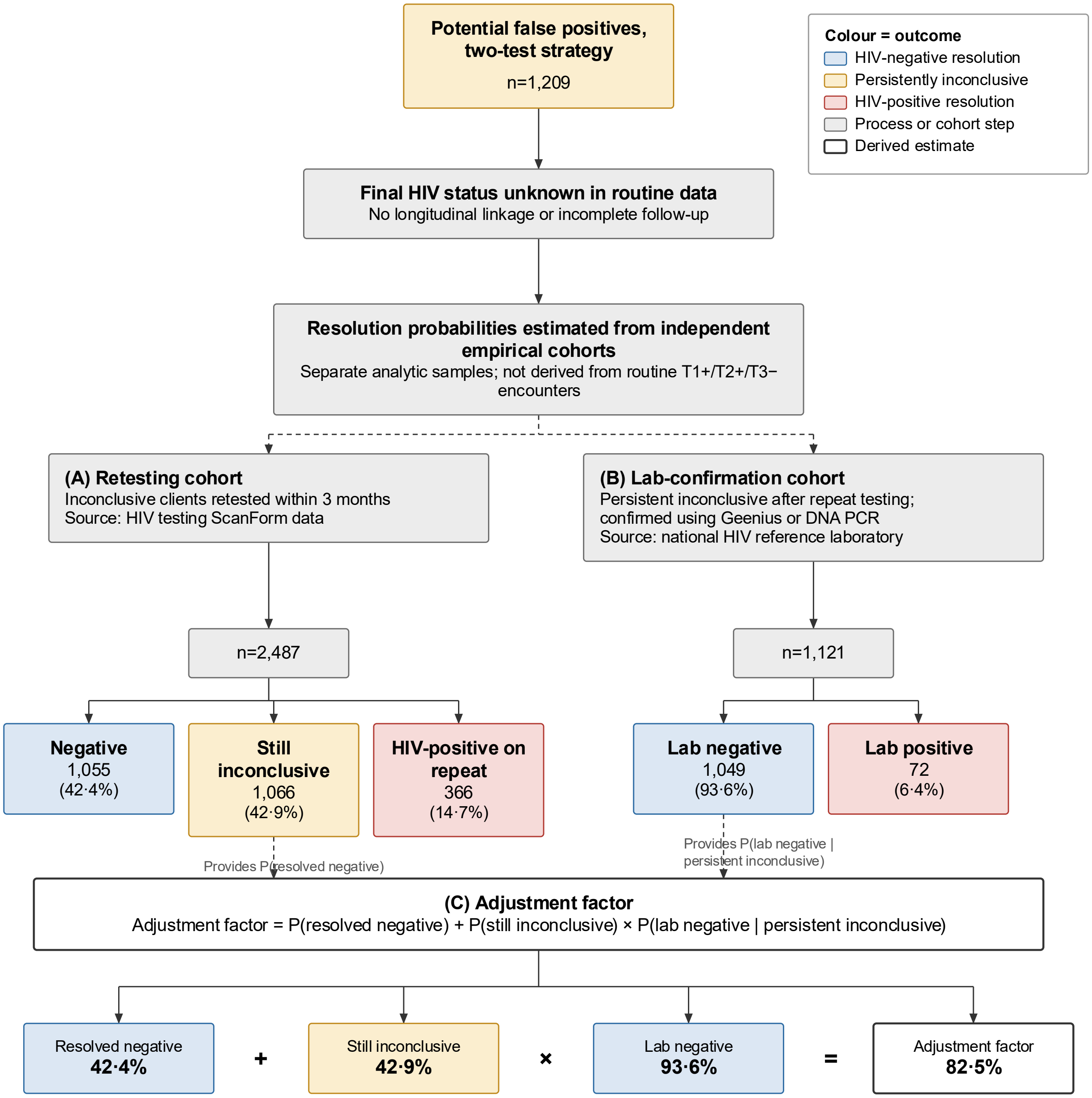

**Figure S3** Resolution outcome flow for T1+/T2+/T3− encounters

Pathways by which inconclusive encounters were ultimately classified as HIV-negative or HIV-positive, based on HTS retesting cohort and national reference laboratory data.

#### Appendix S3. Metric definitions and derivations

The simulated two-test classification is cross-tabulated against the three-test reference.

**Table S3**-Two-by-two classification matrix comparing simulated two-test and three-test classifications

| Simulated two-test classification | Three-test positive (T1+/T2+/T3+) | Three-test non-positive (negative or inconclusive) | Total |
|---|---|---|---|
| Positive after T1 and T2 | a = 171,351 | b = 1,209 | a+b = 172,560 |
| Non-positive | c = 0* | d = 9,690,348 | c+d = 9,690,348 |
| **Total** | **a+c = 171,351** | **b+d = 9,691,557** | **N = 9,862,908** |

**a** = T1+/T2+/T3+, confirmed positive by the three-test strategy; **b** = T1+/T2+/T3−, reactive after T1 and T2 but not confirmed by T3; **c** = three-test positive but simulated two-test non-positive; **d** = non-positive under both strategies.

*Cell c is structurally zero because a three-test positive classification requires reactivity at T1 and T2.

**Table S4** Definitions, formulas, pathways, substitutions, and reported estimates for study metrics

| Metric and definition | Formula | Numerator pathway | Denominator pathway | Substitution and reported estimate |
|---|---|---|---|---|
| **Positive predictive value (PPV)** Proportion of T1+/T2+ encounters confirmed positive by T3. | PPV = a/(a+b) | T1+ → T2+ → T3+ a = 171,351 | All T1+ → T2+ encounters, whether T3+ or T3− a+b = 172,560 | 171,351/172,560 × 100 = 99·30% 95% CI 99·26–99·34 |
| **Potential false-positive proportion (1−PPV)** Proportion of T1+/T2+ encounters not confirmed by T3. | 1−PPV = b/(a+b) | T1+ → T2+ → T3− b = 1,209 | All T1+ → T2+ encounters, whether T3+ or T3− a+b = 172,560 | 1,209/172,560 × 100 = 0·70% 95% CI 0·66–0·74 |
| **Apparent Specificity** Proportion of three-test negative or inconclusive encounters also non-positive under the simulated two-test strategy. | Specificity = d/(b+d) | Non-positive under both strategies d = 9,690,348 | All three-test negative or inconclusive encounters b+d = 9,691,557 | 9,690,348/9,691,557 × 100 = 99·99% 95% CI 99·99–99·99 |
| **Potential false-positive frequency per 100 000 three-test non-positives** Number of potential false-positive classifications per 100 000 three-test negative or inconclusive encounters. | [b/(b+d)] × 100 000 | T1+ → T2+ → T3− b = 1,209 | All three-test negative or inconclusive encounters b+d = 9,691,557 | 1,209/9,691,557 × 100 000 = 12·5 per 100 000 95% CI 11·8–13·2 |

### Appendix S4. Validation of the estimation approach

#### S4.1 Stratified analyses

To assess whether the probability of HIV-negative classification differed across demographic groups, estimates were calculated separately by sex and age group using stratified HTS retesting and laboratory datasets. Estimates were similar for males and females (0·82 and 0·83, respectively); across age groups, estimates ranged from 0·81 to 0·84. Because variation across strata was minimal, the overall estimate (0·825) was applied in the primary analysis. Age-stratified results are shown in **figure S4**.

#### S4.2 Facility-level validation

Facility-level counts of expected persistent inconclusive cases (derived from HTS data) and samples processed by the national reference laboratory showed a strong positive correlation, indicating that laboratory datasets captured the majority of persistent inconclusive referrals. The facility-level comparison is shown in **figure S5**.

#### S4.3 Processing ratio analysis

Facility-level processing ratios (processed laboratory samples divided by expected persistent inconclusive samples) were close to 1·0 at most facilities, indicating high completeness of laboratory referral. Distributions are shown in **figure S6**.

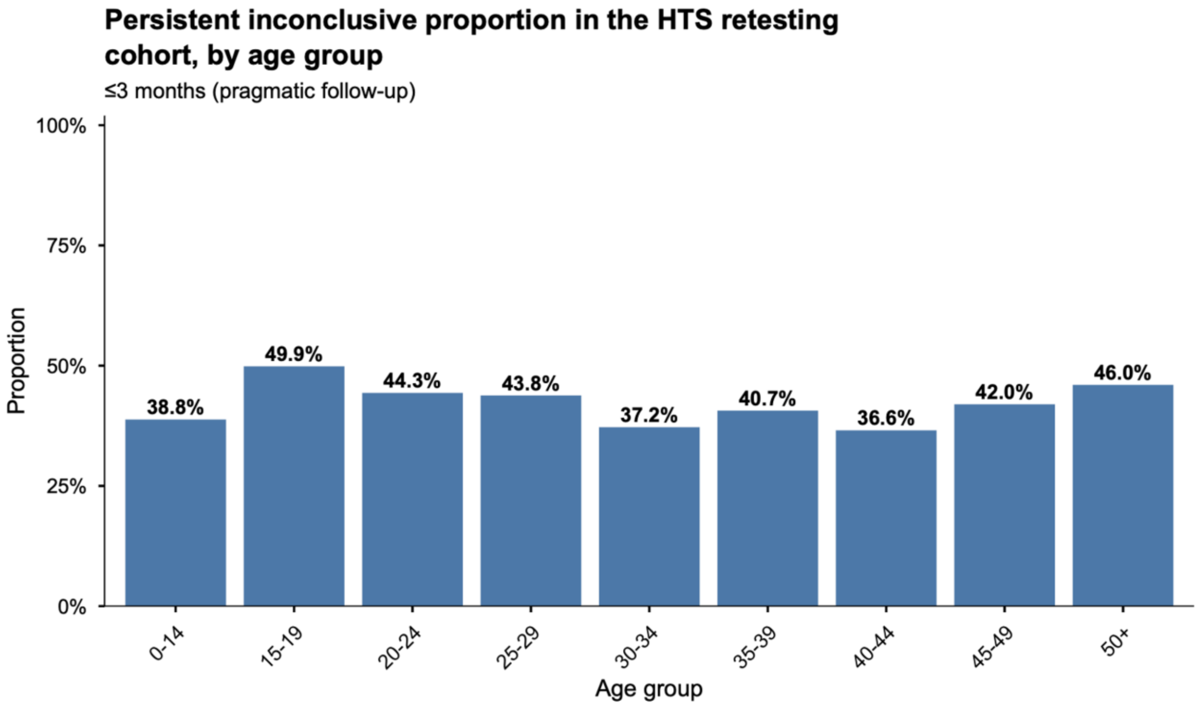

**Figure S 4.** Persistent inconclusive outcomes in the HTS retesting cohort, by age group

Percentage of repeat testing encounters among previously inconclusive clients that remained persistently inconclusive, stratified by age group. Outcomes were broadly consistent across age groups.

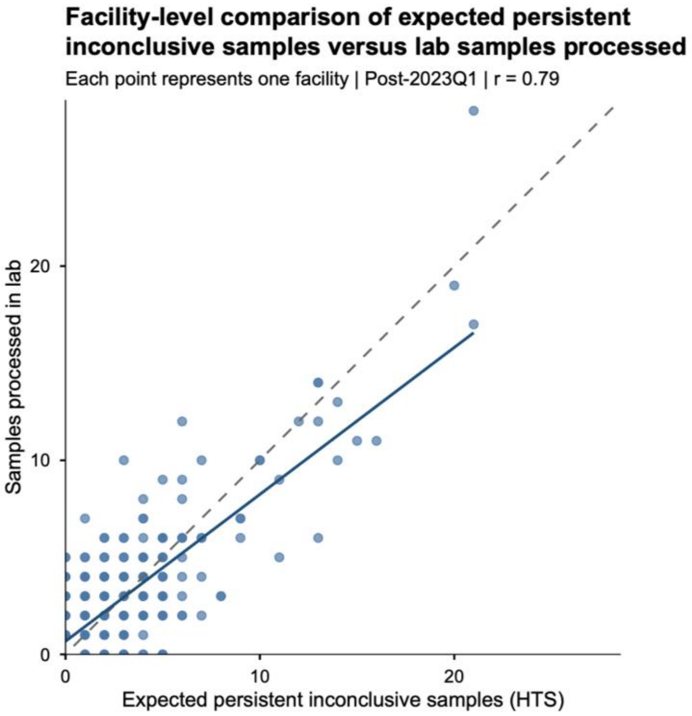

**Figure S5.** Facility-level comparison of expected persistent inconclusive cases and laboratory processed samples
Relationship between facility-level counts of persistent inconclusive cases identified in HTS data and samples processed by the national reference laboratory. Each point represents one facility. The strong positive correlation supports completeness of the laboratory dataset and that, despite individual record linkage not being possible, in aggregate the laboratory dataset likely accurately reflects adjudicated HIV status among individuals referred for lab confirmation following repeated inconclusive testing encounters.

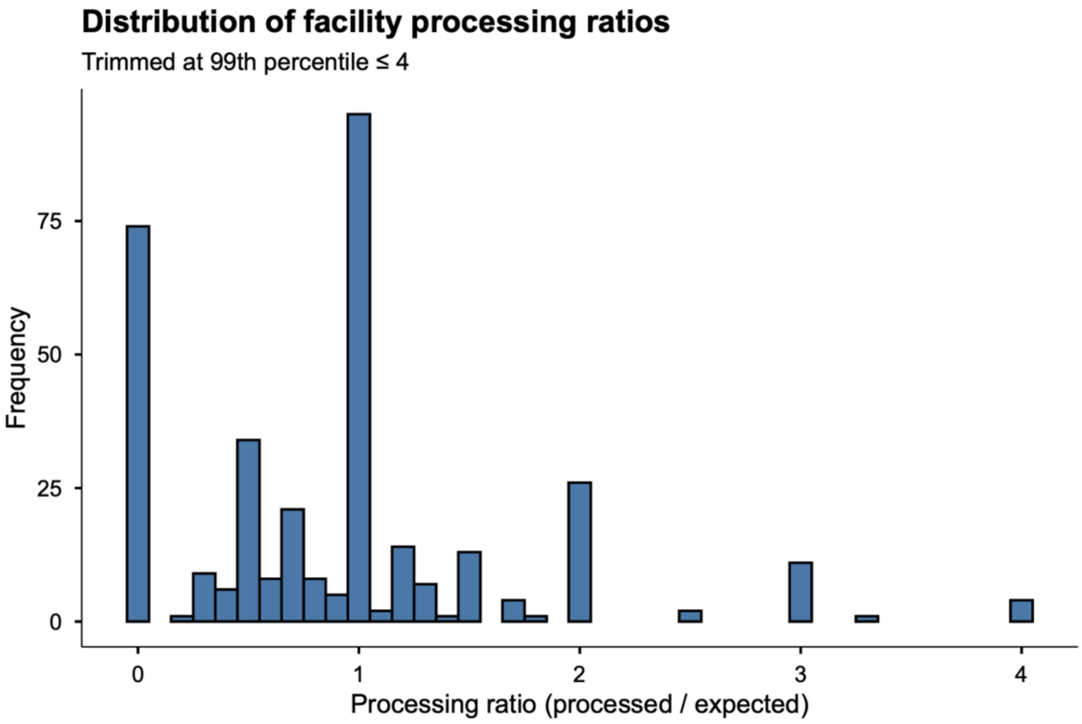

**Figure S 6.** Distribution of facility-level processing ratios

Distribution of facility-level processing ratios (processed laboratory samples divided by expected persistent inconclusive cases). Ratios trimmed at the 99th percentile to reduce the influence of extreme values.

### Appendix S5. Costing methods and sources

#### S5.1 Costing approach

We compared the cost of implementing the three-test algorithm with a counterfactual two-test algorithm from a health-system payer perspective.[4] The comparison combines diagnostic testing costs (commodities and personnel) with avoided lifetime ART care costs (commodities, personnel, and delivery overhead). Assessment of both HIV testing costs and lifetime ART costs include commodities, service delivery personnel, and delivery management overhead, supporting a like-for-like comparison. All costs are expressed in 2025 US dollars.

#### S5.2 Diagnostic commodity costs

Unit commodity costs for the three rapid diagnostic tests were taken from the Global Fund pooled procurement reference prices for 2025:[5]

**Table S5**- Diagnostic commodity costs

| Test | Manufacturer | Pack size | Unit cost (USD) |
|---|---|---|---|
| T1 − Determine HIV-1/2 | Abbott Diagnostics | 100 | $0·80 |
| T2 − Uni-Gold HIV | Trinity Biotech | 20 | $1·60 |
| T3 − SD Bioline HIV-1/2 3.0 | Standard Diagnostics | 25 | $0·80 |

These prices were applied to all kits used under each algorithm, including additional kit requirements for quality control, training, losses, and expiry. Programme wastage rates of 4% for T1 and 43% for T2 and T3 were derived from Malawi programme data. The much higher rates of wastage for T2 and T3 reflect two things: a larger share of these kits is used for training and quality assurance rather than client testing, and each site uses far fewer T2 and T3 kits because only a small proportion of clients are T1-reactive (on average about 60 T1 kits are used for every T2 or T3 kit). At these low volumes a larger fraction of stock is lost to expiry and to partial-pack discard before it can be used, so the effective wastage rate is high even though the absolute number of T2 and T3 kits is small.

#### S5.3 Diagnostic personnel costs

Personnel costs were derived from a published Malawi HIV testing-episode cost (US$2·85 per episode in 2025 USD).[6] The personnel share (50%) corresponds to US$1·43 per episode and represents health-worker time per testing encounter. In the main analysis we did not restrict the incremental non-commodity cost to personnel alone: we applied the full non-kit cost of US$1·955 per incremental client-use testing step (US$1·425 personnel, US$0·202 supervision and monitoring and evaluation, and US$0·328 other delivery and overhead; table S6), so that each additional testing step carries the same delivery, management, and overhead loading as a routine testing encounter.

**Table S 6** Components of the published Malawi HIV testing-episode cost (US$2·85 per episode)

| Component | Share of episode cost | Cost per episode (USD) | Grouped category |
|---|---|---|---|
| Personnel or health-worker time | 50·0% | $1·43 | Delivery |
| HIV test kits | 31·4% | $0·89 | Commodity |
| National or district supervision, audits and M&E | 7·1% | $0·20 | Management |
| Other supplies, utilities, capital, overheads | 11·5% | $0·33 | Other delivery or overhead |
| **Total** | **100·0%** | **$2·85** | |

Personnel cost was applied only to the incremental client-use testing steps generated by the three-test algorithm beyond what the two-test counterfactual would have required, not to every assay performed. Applying personnel cost to every assay would over-counted fixed encounter-level costs shared between the two algorithms. An incremental client-use testing step was defined as any additional T2 or T3 assay performed under the three-test algorithm that would not have been performed under the two-test counterfactual.

##### S5·4 Two-test counterfactual

For the two-test counterfactual, discordant T1+/T2− results were assumed to undergo same-session repeat parallel testing in accordance with Malawi's national HIV testing guidelines.[3] The proportion of discordant results remaining discordant after this same-session repeat was set at 40%, derived from aggregate Department of HIV and AIDS Management Information System (DHAMIS) data covering the two-test implementation period before November 2022. Encounters remaining discordant after same-session repeat were assumed to require follow-up testing under the simulated two-test strategy.

##### S4·5 ART care costs avoided

For each false-positive diagnosis averted, we estimated the lifetime ART care costs that would have been incurred had the diagnosis stood.[7] The annual ART care cost was estimated at US$72·87 per patient-year, rounded to US$73 in the main text. The cost was built up by replacing the commodity components of a published rural Malawi provider-costing study[6] with current Malawi Global Fund forecast and quantification figures,[5] while retaining the delivery, management, and overhead components from the costing study.

**Table S 7** Components of the Malawi-adapted annual ART care cost (US$72·87 per patient-year)

| Component | Cost per patient-year (USD) | Source | Grouped category |
|---|---|---|---|
| ART drugs (ARV commodities) | $40·33 | Malawi Global Fund forecast | Commodity |
| Cotrimoxazole | $0·00 | Included within ARV figure | Commodity |
| Viral-load testing | $14·76 | Malawi Global Fund forecast | Laboratory monitoring |
| Personnel | $10·79 | Rural Malawi provider-costing study | Delivery |
| Capital inputs or training | $4·06 | Rural Malawi provider-costing study | Delivery |
| District or national supervision | $1·77 | Rural Malawi provider-costing study | Management |
| Other supplies, utilities and overheads | $1·16 | Rural Malawi provider-costing study | Other delivery or overhead |
| **Total** | **$72·87** | | |

For reference, the unadjusted published rural Malawi provider-costing study[6] estimated US$99·35 per patient-year (ART drugs $70·73, cotrimoxazole $5·89, VL testing $4·50, personnel $10·79, capital or training $4·06, supervision $1·77, other $1·61). Substituting current Global Fund commodity prices reduces the commodity components and increases the VL component (reflecting the substantial scale-up of viral-load coverage since the costing study), while leaving the delivery and management components unchanged.

##### S5.6 Discounting and time horizon

Avoided lifetime ART care costs were calculated over a 30-year time horizon and discounted at 3% per annum. The 30-year horizon corresponds to the modal life expectancy on ART in sub-Saharan African settings from a typical age of diagnosis and is consistent with prior cost-effectiveness analyses of HIV testing in the region.

#### S5.7 Sensitivity analyses

We performed sensitivity analyses varying:

- Kit overhead not used for direct patient testing (±20% around the observed values, applied multiplicatively to the base rates of 4% for T1 and 43% for T2 and T3). These rates represent kits used for quality control, training, losses, and expiry rather than direct patient testing.
- Persistence of T1+/T2− discordance after same-session repeat testing under the simulated two-test strategy (25% to 75%, around the observed value of 40%). This assumption affects the amount of additional follow-up testing required under the two-test counterfactual and, consequently, the incremental cost of transitioning to the three-test strategy.
- Diagnostic costing scenario. The main analysis applied the full non-kit cost of US$1·955 per additional client-use testing step (rounded to US$1·96), comprising personnel, supervision and monitoring and evaluation, and other delivery and overhead costs. We compared this with two lower-cost alternatives under the base-case wastage and discordance assumptions: a commodity-only scenario and a commodity-plus-personnel scenario.

The combined sensitivity analysis for kit overhead and persistent discordance under the main full non-kit costing scenario is presented in figure S 7. Incremental diagnostic testing costs ranged from US$437,679 to US$489,887, with corresponding discounted times to cost offset ranging from 6·74 to 7·65 years. Across all scenarios displayed in figure S 7, discounted ART care costs avoided (US$1,424,606) exceeded incremental diagnostic testing costs.

#### S5.8 Costs not included

We did not estimate costs from the patient or societal perspective. These would include time and transport costs of testing encounters, indirect economic costs of false-positive diagnosis (lost earnings, household disruption), and psychosocial costs (anxiety, stigma, partnership effects). The true societal cost of misdiagnosis is therefore higher than the provider-perspective cost reported here. We also did not include training costs associated with the transition to the three-test algorithm; as a one-time investment delivering benefits over many years, this omission is unlikely to alter the cost-saving conclusion.

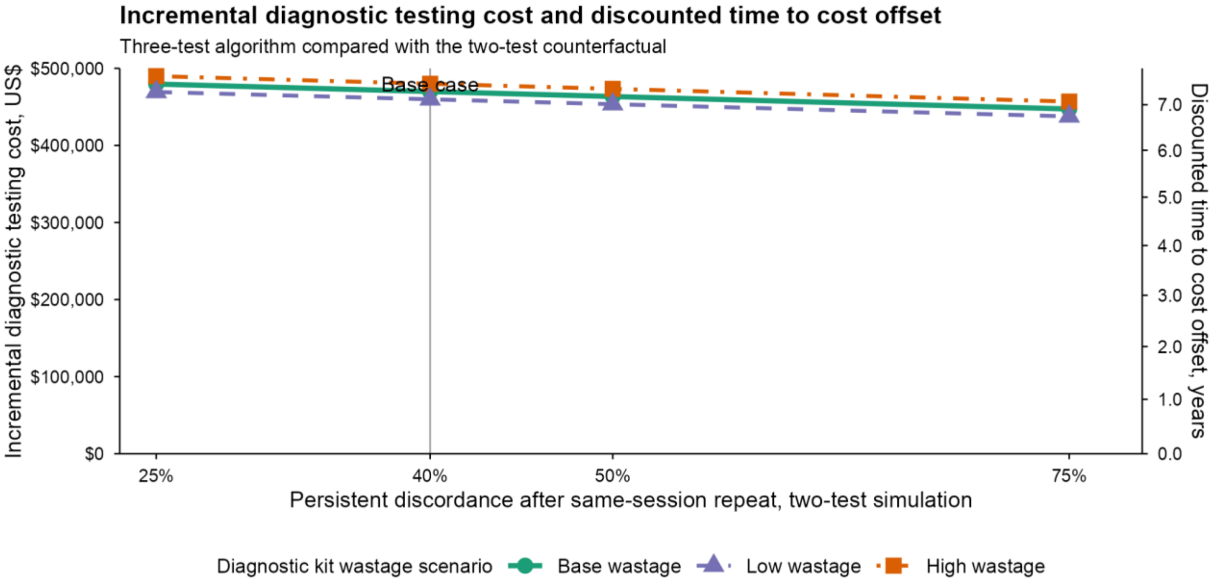

**Figure S 7** Incremental diagnostic commodity cost of the three-test algorithm sensitivity scenarios

#### Appendix S6. STARD checklist

| Section & Topic | No | Item | Reported on page # |
|---|---|---|---|
| **TITLE OR ABSTRACT** | | | |
| | 1 | Identification as a study of diagnostic accuracy using at least one measure of accuracy (such as sensitivity, specificity, predictive values, or AUC) | p. 1 (title); p. 2 (Abstract — Findings) |
| **ABSTRACT** | | | |
| | 2 | Structured summary of study design, methods, results, and conclusions (for specific guidance, see STARD for Abstracts) | pp. 2–3 (Abstract) |
| **INTRODUCTION** | | | |
| | 3 | Scientific and clinical background, including the intended use and clinical role of the index test | p. 5 (Introduction, paragraph 1) |
| | 4 | Study objectives and hypotheses | p. 6 (Introduction, paragraph 2, final sentence) |
| **METHODS** | | | |
| Study design | 5 | Whether data collection was planned before the index test and reference standard were performed (prospective study) or after (retrospective study) | p. 1 (title); pp. 6–7 (Methods — Study design, setting, and data sources) |
| Participants | 6 | Eligibility criteria | p. 7 (Methods — Inclusion and exclusion) |
| | 7 | On what basis potentially eligible participants were identified (such as symptoms, results from previous tests, inclusion in registry) | p. 7 (Methods — Inclusion and exclusion) |
| | 8 | Where and when potentially eligible participants were identified (setting, location and dates) | pp. 6–7 (Methods — Study design, setting, and data sources); p. 11 (Results, opening paragraph) |
| | 9 | Whether participants formed a consecutive, random or convenience series | p. 7 (Methods — Inclusion and exclusion) |
| Test methods | 10a | Index test, in sufficient detail to allow replication | pp. 7–8 (Methods — HIV diagnostic test algorithm and analytic approach, paragraphs 1–2) |
| | 10b | Reference standard, in sufficient detail to allow replication | pp. 7–8 (Methods — HIV diagnostic test algorithm and analytic approach, paragraph 1) |
| | 11 | Rationale for choosing the reference standard (if alternatives exist) | p. 8 (Methods — HIV diagnostic test algorithm and analytic approach); p. 17 (Discussion — limitations paragraph) |

| Section & Topic | No | Item | Reported on page # |
|---|---|---|---|
| | 12a | Definition of and rationale for test positivity cut-offs or result categories of the index test, distinguishing pre-specified from exploratory | p. 8 (Methods — HIV diagnostic test algorithm and analytic approach, paragraph 2) |
| | 12b | Definition of and rationale for test positivity cut-offs or result categories of the reference standard, distinguishing pre-specified from exploratory | pp. 7–8 (Methods — HIV diagnostic test algorithm and analytic approach, paragraph 1) |
| | 13a | Whether clinical information and reference standard results were available to the performers/readers of the index test | Not applicable — index test simulated post hoc from recorded test sequence (see Methods, p. 8) |
| | 13b | Whether clinical information and index test results were available to the assessors of the reference standard | Not applicable — reference standard is sequential by protocol (see Methods, pp. 7–8) |
| Analysis | 14 | Methods for estimating or comparing measures of diagnostic accuracy | pp. 8–9 (Methods — HIV diagnostic test algorithm and analytic approach, paragraph 4) |
| | 15 | How indeterminate index test or reference standard results were handled | p. 8 (Methods — HIV diagnostic test algorithm and analytic approach, paragraph 2); p. 10 (Methods — Estimating the proportion of false-positive results among inconclusive results) |
| | 16 | How missing data on the index test and reference standard were handled | p. 9 (Methods — Factors associated with a potential false-positive diagnosis, final sentence) |
| | 17 | Any analyses of variability in diagnostic accuracy, distinguishing pre-specified from exploratory | p. 9 (Methods — Factors associated with a potential false-positive diagnosis); pp. 10–11 (Methods — Diagnostic commodity cost analysis, sensitivity analyses) |
| | 18 | Intended sample size and how it was determined | pp. 6–7 (Methods — Study design, setting, and data sources) |
| **RESULTS** | | | |
| Participants | 19 | Flow of participants, using a diagram | Figure 1 (provided separately) |
| | 20 | Baseline demographic and clinical characteristics of participants | pp. 12–13 (Results, final paragraph before Diagnostic performance); Table 2 (provided separately) |
| | 21a | Distribution of severity of disease in those with the target condition | Not applicable — HIV diagnosis is not graded by severity in this context |
| | 21b | Distribution of alternative diagnoses in those without the target condition | Not applicable — alternative diagnoses not recorded in routine testing registers; biological mechanisms discussed pp. 16 and 18 (Discussion) |
| | 22 | Time interval and any clinical interventions between index test and reference standard | p. 8 (Methods — HIV diagnostic test algorithm and analytic approach); p. 10 (Methods — Estimating the proportion of false-positive results among inconclusive results) |

| Section & Topic | No | Item | Reported on page # |
|---|---|---|---|
| Test results | 23 | Cross tabulation of the index test results (or their distribution) by the results of the reference standard | p. 13 (Results); Table 1 (provided separately) |
| | 24 | Estimates of diagnostic accuracy and their precision (such as 95% confidence intervals) | pp. 13–14 (Results); Tables 1 and 2 (provided separately) |
| | 25 | Any adverse events from performing the index test or the reference standard | Not applicable — finger-prick capillary HIV RDTs are minimally invasive; no adverse events recorded or expected |
| **DISCUSSION** | | | |
| | 26 | Study limitations, including sources of potential bias, statistical uncertainty, and generalisability | p. 17 (Discussion — limitations paragraph); generalisability discussed pp. 16 and 18 |
| | 27 | Implications for practice, including the intended use and clinical role of the index test | pp. 17–18 (Discussion, concluding paragraphs) |
| **OTHER INFORMATION** | | | |
| | 28 | Registration number and name of registry | Not prospectively registered (retrospective analysis of routine programme data); ethical approval p. 11 (Methods — Ethical approval; Malawi NHSRC Protocol 23/12/4275) |
| | 29 | Where the full study protocol can be accessed | p. 19 (Data sharing) — statistical analysis plan available from the corresponding author on request |
| | 30 | Sources of funding and other support; role of funders | p. 11 (Methods — Role of the funding source); p. 19 (Acknowledgements) |

#### References

**1.** Ministry of Health Malawi. HIV, syphilis, and hepatitis B integrated rapid testing guidelines and operating procedures. 1st ed. Lilongwe: Ministry of Health; 2023.

**2** World Health Organization. Consolidated guidelines on HIV testing services, 2019. Geneva: WHO; 2019.

**3.** Ministry of Health Malawi. HIV testing services guidelines. Lilongwe: Ministry of Health; 2016.

**4.** Drummond MF, Sculpher MJ, Claxton K, et al. Methods for the economic evaluation of health care programmes. 4th ed. Oxford: Oxford University Press; 2015.

**5.** The Global Fund. Pooled procurement mechanism reference pricing: RDTs. Geneva: The Global Fund; 2025.

**6.** Vyas S, Songo J, Guinness L, et al. Assessing the costs and efficiency of HIV testing and treatment services in rural Malawi: implications for future 'test and start' strategies. BMC Health Serv Res 2020; 20: 740.

**7.** Eaton JW, Johnson CC, Gregson S. The cost of not retesting: human immunodeficiency virus misdiagnosis in the antiretroviral therapy test-and-offer era. Clin Infect Dis 2017; 65: 522–25.
